# Percutaneous Spinal Stimulation to Enhance Gait and Alleviate Functional Impairments in Multiple Sclerosis: A Pilot Study

**DOI:** 10.1101/2025.06.23.25329591

**Authors:** Omid Jahanian, Anders J. Asp, Megan L. Gill, Daniel D. Veith, K. A. Fernandez, Cecilia A. Hogen, Candee J. Mills, Andrew R. Thoreson, Ryan Solinsky, Kenton R. Kaufman, Peter J. Grahn, Kristin D. Zhao, W. Oliver Tobin

## Abstract

**Objective:** To evaluate the feasibility and potential efficacy of percutaneous spinal stimulation (epidural stimulation, ES) combined with task-specific training to reduce spasticity and improve gait and balance in individuals with progressive multiple sclerosis (MS).

**Methods:** Two men with progressive MS (EDSS 6.5) underwent ES lead implantation targeting the lower spinal cord, followed by one month of rehabilitation involving 12 ES-assisted training sessions. Assessments included instrumented gait analysis, Modified Ashworth Scale (MAS), pendulum test, and static balance testing performed at baseline, and at end of study with ES-Off and ES-On conditions.

**Results:** Participant 1, with spastic hemiparetic gait, demonstrated improved lower extremity joint kinematics and muscle activation during gait, reduced knee extensor spasticity, and enhanced functional movement patterns with ES-On. Participant 2, with significant paraparesis and minimal spasticity, showed limited gait changes but experienced marked improvements in static balance, particularly under eyes-closed conditions. No adverse events were reported.

**Conclusion:** This pilot study suggests that ES paired with task-specific training may enhance gait and balance in individuals with progressive MS. Responses varied based on impairment profiles, underscoring the importance of individualized ES parameter tuning. These findings warrant further investigation of ES as a therapeutic option for motor impairments in MS.

## INTRODUCTION

Progressive motor impairment, specifically myelopathy, is the most common presentation of progressive multiple sclerosis (MS).^1, 2^ Mounting evidence suggests that cortical spinal tract involvement with MS lesions is responsible for the development of motor disability, primarily characterized by gait dysfunction.^3–6^ Gait issues, often associated with spasticity,^7, 8^ weakness,^9^ and impaired balance,^10^ are among the most debilitating MS symptoms, profoundly affecting daily function and quality of life.^11^

Up to 75% of patients with MS develop progressive disease, with no current treatment to halt or reverse its disability. Pathologically, progressive MS is characterized by gradual axonal loss in the absence of overt inflammatory disease. Current treatment options are limited to addressing the inflammatory component of the disease; they do not halt or reverse the neuronal death and axonal loss that contribute to declining motor function and walking ability. Progressive motor dysfunction in MS is most common in patients who have spinal cord lesions or lesions in the brainstem involving the motor tracts.^5, 12, 13^ Complete loss of axons in any particular tract, even in patients with progressive MS, is unusual. Spinal cord stimulation, which is believed to activate remaining neuronal pathways, has shown potential for facilitating volitional motor function, as demonstrated in individuals with spinal cord injury.^14–18^ However, its potential as a promising intervention to address motor dysfunction in MS patients is understudied.

Spinal cord stimulation, particularly epidural spinal cord stimulation (ES), was initially developed in the 1960s to treat chronic pain by suppressing noxious input. ^19^ Unexpectedly, ES was discovered to enable motor function in an MS patient, where it was originally intended to relieve pain.^20^ Despite these early findings, the potential of ES to enhance motor function in MS patients remained largely unexplored for many years. In the following decades, ES research focused on determining effective stimulation parameters to improve motor function, specifically in spinal cord injury,^21^ where ES has been shown to enhance volitional motor function and improve seated posture, reaching, standing, and stepping.^14–16, 18^

Deficits following MS can be highly individualized, and one of the key benefits of ES over traditional approaches to improve gait function, is that it can be tailored to an individual’s disability to address specific impairments, such as weakness or spasticity. This highlights the need for further investigation into the effects of ES on progressive MS to strengthen the evidence base for clinical application. This study aims to investigate the feasibility and potential efficacy of temporary ES to reduce spasticity, while improving balance and gait in individuals with progressive MS.

## METHODS

### Study Participants

Following FDA investigational device exemption and Mayo Clinic Institutional Review Board approval, two male participants with progressive MS met inclusion criteria and were enrolled in this pilot study (NCT06019611). Inclusion criteria included no clinical or radiologic MS relapses for over 5 years, an Expanded Disability Status Scale (EDSS) score of 6.5, the ability to ambulate 10 feet independently with or without a gait aid, and no changes to spasticity medications or dalfampridine in the last 3 months. Exclusion criteria included Grade 4 spasticity measured bilaterally in four muscle groups using the Modified Ashworth Scale (MAS), active participation in an interventional clinical trial, any active implanted medical device, and any illness or condition that would compromise the participant’s ability to comply with the protocol or their safety.

### Study Design

As outlined in Figure 1, baseline assessments, including gait and balance analysis (Figure 2A), were obtained prior to ES lead implantation, described as No ES. One day after implantation, lead location and potential motor activation was determined by performing spatial temporal electrophysiological mapping of the lumbosacral spinal cord, iterating through multiple electrode combinations while adjusting stimulation amplitude (mA) and frequency (Hz).^22^ Subsequently, the participants completed one month of rehabilitation, which included ES and task-specific training across 12, two-hour sessions. Finally, prior to explanting the leads, each participant underwent post-rehabilitation assessments, including spasticity assessment, and gait and balance analysis, with stimulation off (ES-Off) and on (ES-On), (Figure 2F).

**Figure 1.**
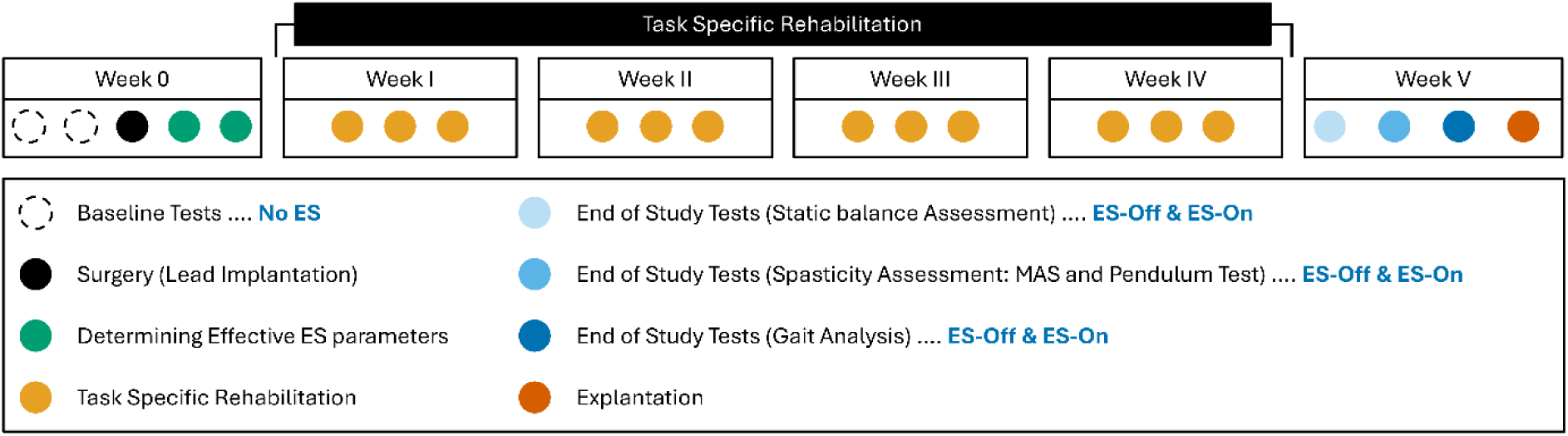
Methodological Approach.

**Figure 2.**
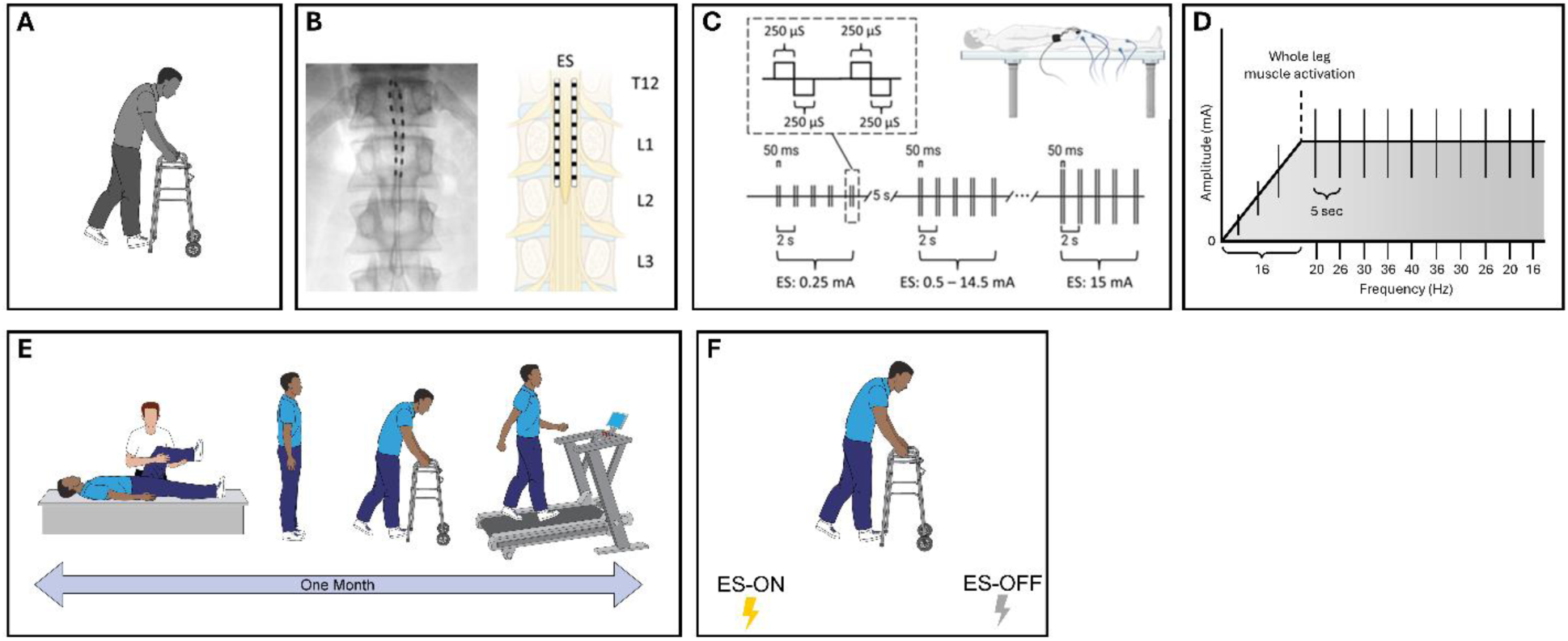
Study design. A. Baseline tests. B. Fluoroscopic image and illustration of ES lead placement (created with BioRender). C. Motor recruitment curves to enable motor activation of the lower extremities. D. Frequency curves. E. ES and task-specific training, and F. End of study test with ES-On and ES-Off.

### Implantation Surgery

Two temporary percutaneous epidural stimulation leads (Abbott Neuromodulation, Plano, TX) were placed parallel to one another along the dorsal epidural surface of the lumbosacral enlargement (Figure 2B). The externalized ends of the ES electrode leads were connected to a neurostimulation system (Abbott programmer; Abbott, Plano, TX) for delivering electrical pulse waveforms to the epidural surface. To refine electrode array alignment, intraoperative myography was used to record ES-evoked motor potentials from LE muscles bilaterally^23^.

### Spatial Temporal Electrophysiological Mapping of the Lumbosacral Spinal Cord

To confirm lead location and determine the general muscle activation patterns of the LE’s, baseline motor recruitment curves were collected on day one of testing following surgical implantation (Figure 2C). Motor recruitment curves were generated with a train of five charge-balanced, biphasic paired pulses each with a pulse width of 250 µs and separated by 50ms with different electrode configurations. Amplitude was increased stepwise (0.25-15.0mA) at a frequency of 0.5Hz to enable motor activation of the lower extremities (Ripple Neuromed, Salt Lake City, UT, USA). To measure evoked electromyography (EMG) responses specific to lower-limb muscles, electrodes were connected to a 16-channel biosignal amplifier (g.BSamp, g.tec medical engineering GmbH, Schiedlberg, Oberösterreich, Austria). Each channel was configured with a gain of 100 μV/V. The resulting surface EMG signals were amplified and digitized at a sampling rate of 4 kHz using a PowerLab 16/35 system (AD Instruments, Dunedin, New Zealand). Motor activation thresholds were defined as LE EMG recordings exceeding 50µv and three standard deviations beyond resting motor activity while in the supine position.^22^ On day 2, frequency curves were collected at the lowest amplitude of whole leg muscle activation identified through EMG, with similar stepwise progression from 16 to 40Hz (Figure 2D).

Recruitment and frequency curves were collected in a passive, supine position. The LE muscles studied to determine proximal versus distal activation patterns include the rectus femoris, vastus lateralis, medial hamstring, gastrocnemius, soleus, and tibial anterior muscle groups.

### ES and Task-Specific Training

The participants completed one month of physical therapy, which included ES paired with task-specific training across 12 two-hour sessions (Figure 2E). ES was applied using information obtained during the spatial temporal electrophysiological mapping including muscle activation specificity based on electrode location (proximal vs. distal). Stimulation parameters were fine-tuned at each session to enhance motor performance during task-specific training and were used as the effective ES parameters for the end-of-study tests. The task-specific training consisted of repeated gait activities on both the treadmill and overground, with or without body weight support, upright standing and balance exercises, pre-gait activities, and isolated strength training of the LE muscles. All task-specific training activities were done with stimulation on (ES-ON) and stimulation was only available when the participant was in the laboratory. Rest breaks from both the task-specific training and stimulation were provided intermittently based on performance and/or the participant’s request.

### Data Collection & Processing

#### Modified Ashworth Scale (MAS)

Spasticity was assessed using the MAS with participants positioned in supine with a 10-inch therapy wedge supporting their trunk while their legs hung off the edge of the bed. Knee flexion/extension and ankle dorsiflexion/plantar flexion were tested by applying a high-velocity stretch, with resistance graded on a 0–4 scale per MAS protocol (Figure 3).^24^

**Figure 3.**
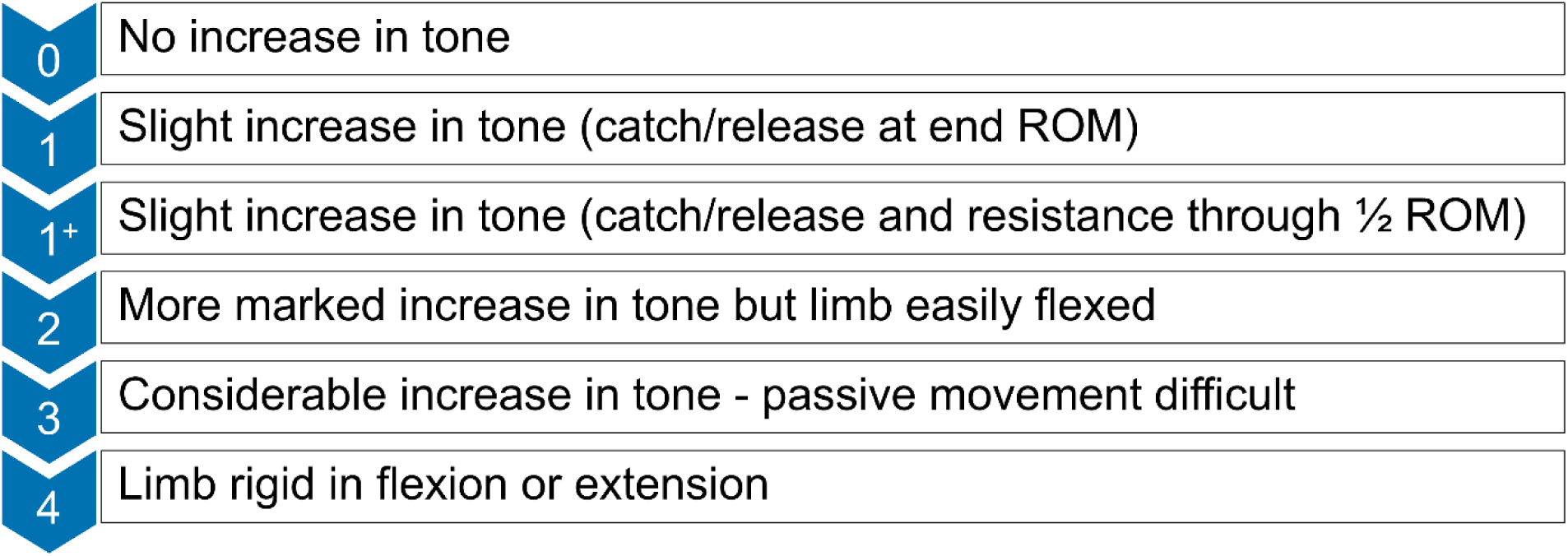
Modified Ashworth Scale Scoring.

#### Pendulum Test

The pendulum test evaluates spasticity by observing the passive swinging of the lower limb under gravity.^25^ The motion resembles a smooth, damped pendulum, while spasticity reduces oscillations and alters the pattern (Figure 4). The pendulum test has good predictive value for detecting the presence of knee extensor muscle spasticity.^26^ In this test, the participants were positioned in the same manner as the MAS; the examiner lifted the relaxed leg to a horizontal position, extended the knee, and released it to swing freely. Three trials per side, with 60-second rests, were recorded using inertial measurement unit (IMU) sensors placed on the shank and thigh (Figure 4). Knee joint angles were derived from shank sensor data, with the first swing angle (FSA; Figure 4) as the primary outcome, correlating with knee extensor spasticity.^27^

**Figure 4.**
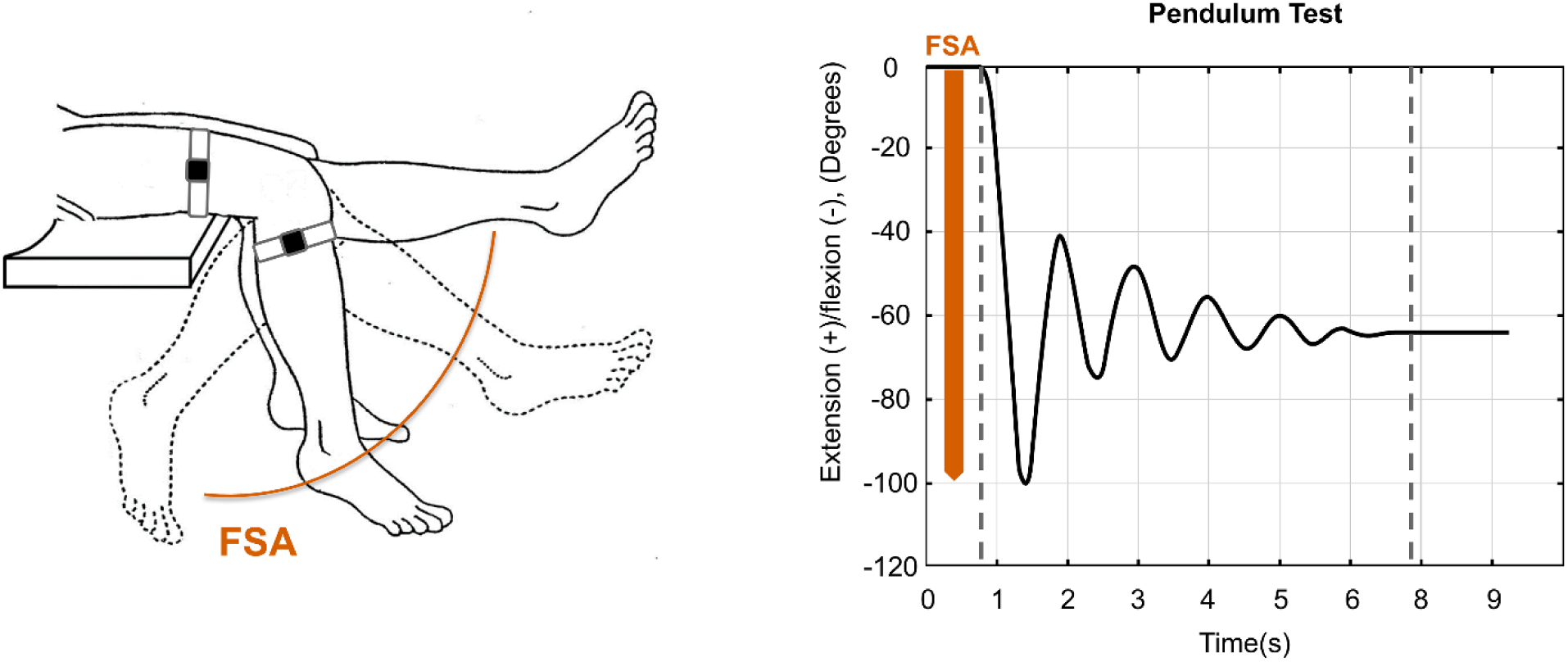
Pendulum test. Kinematic recordings during the pendulum test using an IMU sensor on the shank in an unaffected healthy individual. The first swing angle (FSA; red arrow) was measured as the angle at which the natural backward swing stops its motion below the resting.

#### Gait Analysis

Participants had 41 retro-reflective markers affixed to their extremities, pelvis, trunk, and head. Surface EMG was collected (Motion Lab Systems, Baton Rouge, LA) from LE muscles. Participants walked approximately 10 m at a self-selected, preferred walking speed using a front-wheeled walker (Figure 2F). Marker trajectories of 3–6 strides were recorded by 19 cameras (120 Hz, Raptor-12, Motion Analysis Corporation, Santa Rosa, CA). Kinematic variables were calculated using commercial software (C-Motion Inc., Rockville, MD), with the 3D coordinates of the markers as inputs. Embedded right-hand Cartesian coordinate systems were used to describe the position and orientation of the LE rigid body segments.

Three trials for each of the walking conditions were averaged and used in data analysis for each participant. The period of a gait cycle (stride) was defined from the initial contact of one lower extremity to the next initial contact of that same extremity. All gait variables including the temporospatial parameters (velocity, cadence, step and stride length, double and single support, and step width) and joint kinematics were expressed as a percentage of the gait cycle.

The raw EMG signals were digitized at 4200 Hz, then were full-wave rectified and smoothed using a root mean square (RMS) algorithm with a time-averaging period of 25 ms using a custom program in MATLAB (Mathworks, Natick, MA). The signal magnitude from the RMS envelope of the EMG data was used to determine muscle activity during each gait cycle.

### Postural Control (Static Balance) Assessment

Postural control and static balance was evaluated while standing on an instrumented pressure mat (Tekscan Inc., South Boston, MA), with eyes open (EO) and closed (EC), at baseline with No ES and at the end of study under both ES-Off and ES-On conditions (Figure 1). The participant was asked to stand on the instrumented pressure mat in a quiet stance with arms at their sides and feet positioned approximately hip-width apart. This was repeated in two conditions: EO and EC, with a 60-second resting interval between the tests. The Tekscan system and software were utilized to record the trajectory of the center of force during the balance tests using the instrumented pressure mat and to calculate the balance variables including, sway range in the anterior-posterior and medial-lateral directions (cm), and sway area (cm²). Due to technical issues, static balance tests were performed only on Participant 2.

### Data Analysis

Descriptive statistics including plots and tables for all outcomes were reported for the baseline assessments and end of study assessments under ES-On and ES-Off conditions.

## RESULTS

The two individuals who were enrolled to participate were in their 50s and presented with very different gait impairments. These impairments resemble differences seen across patients in the clinical setting. Participant 1 (male, height: 180 cm, weight: 76 kg) had a primarily spastic hemiparetic gait pattern affecting the right leg greater than the left and presented with a hip circumduction gait pattern. Participant 2 (male, height: 172 cm, weight: 66 kg) had minimal spasticity and had significant paraparesis and presented with a crouched (flexed hips, knees, and ankles) gait pattern, affecting the left leg greater than the right. Both participants utilized a front wheeled walker, could walk 10-20 meters independently with the EDSS of 6.5.

### Baseline Assessments

Gait analysis data during the baseline assessment for Participant 1 indicated a reduced gait speed (0.25 m/s) and decreased range of motion in the LE joints, compared to normative data,^28^ with right knee hyperextension and no ankle dorsiflexion. The data for participant 2 indicated a reduced gait speed (0.38 m/s) and increased ankle dorsiflexion during terminal stance caused by a crouched gait. Table 1 and Figure 5 indicate the spatiotemporal parameters and LE joint kinematics during the baseline assessment.

**Figure 5.**
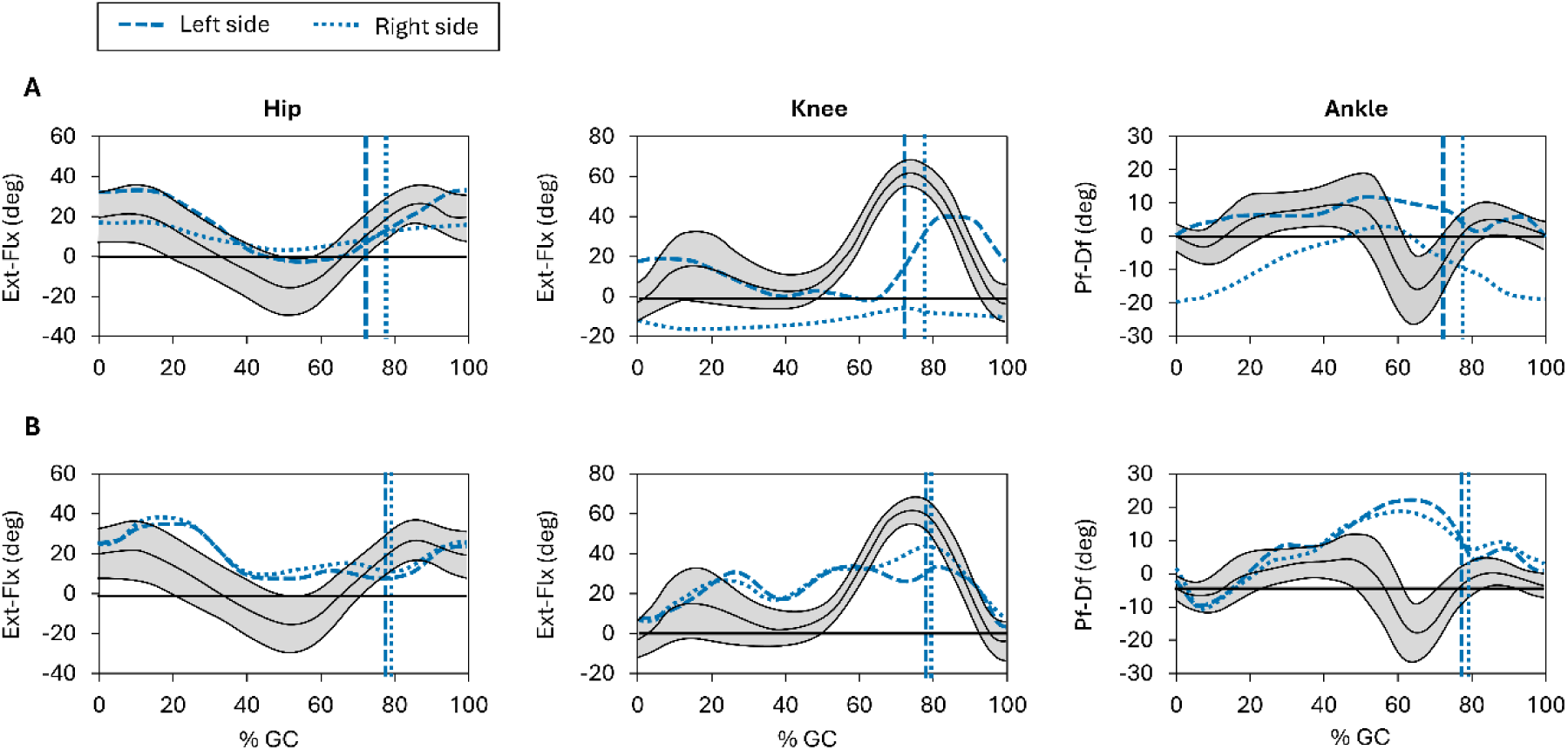
LE joint kinematics in the sagittal plane for participant 1 (A), and Participant 2 (B), during baseline assessment. Joint angles for the hip and knee are expressed as extension–flexion (Ext–Flx), and for the ankle as plantarflexion–dorsiflexion (PF–DF). Each plot displays joint angle trajectories over the gait cycle (%GC; 0–100%). Vertical lines represent transition from stance to swing. Solid black lines represent lab normative data (unpublished), with shaded regions showing ±1 standard deviation. Normative data were collected from 20 healthy adults (10 females; mean (SD): age = 26.7 (4.8) years, height = 173.2 (9.0) cm, mass = 70.1 (12.2) kg, BMI = 23.2 (2.1) kg/m²).

**Table 1.** Spatiotemporal parameters during the baseline and end of study assessments.

|  | Participant 1 |  |  | Participant 2 |  |  | Normative Data |  |  |
| --- | --- | --- | --- | --- | --- | --- | --- | --- | --- |
| Spatiotemporal parameters | Baseline (No Stim) | End of Study, ES-Off | End of Study, ES-On | Baseline (No Stim) | End of Study, ES-Off | End of Study, ES-On | P<sub>5</sub> | Mean | P<sub>95</sub> |
| Velocity (cm/s) | 25 | 17 | 18 | 38 | 31 | 39 | 112 | 138 | 163 |
| Cadence (step/min) | 44 | 29 | 31 | 53 | 50 | 53 | 104 | 120 | 136 |
| Stride (cm) | 70 | 73 | 74 | 87 | 76 | 88 | 120 | 138 | 156 |
| Step Width (cm) | 13 | 11 | 13 | 10 | 12 | 14 | 5 | 10 | 16 |
| Total Support Time (% of cycle) |  |  |  |  |  |  |  |  |  |
| Right | 73 | 77 | 75 | 78 | 80 | 79 | 61 | 63 | 65 |
| Left | 78 | 83 | 85 | 80 | 81 | 80 |  |  |  |
| Single Support Time (% of cycle) |  |  |  |  |  |  |  |  |  |
| Right | 21 | 17 | 15 | 20 | 19 | 21 | 35 | 37 | 39 |
| Left | 28 | 24 | 25 | 22 | 20 | 22 |  |  |  |
| Step Length (cm) |  |  |  |  |  |  |  |  |  |
| Right | 39 | 37 | 37 | 44 | 40 | 45 | 60 | 69 | 78 |
| Left | 31 | 37 | 38 | 43 | 36 | 44 |  |  |  |

Static balance tests at baseline (No ES) with Participant 2 indicated that postural sway was greater during the EC condition compared to the EO condition, particularly in the medial-lateral direction (Figure 6).

**Figure 6.**
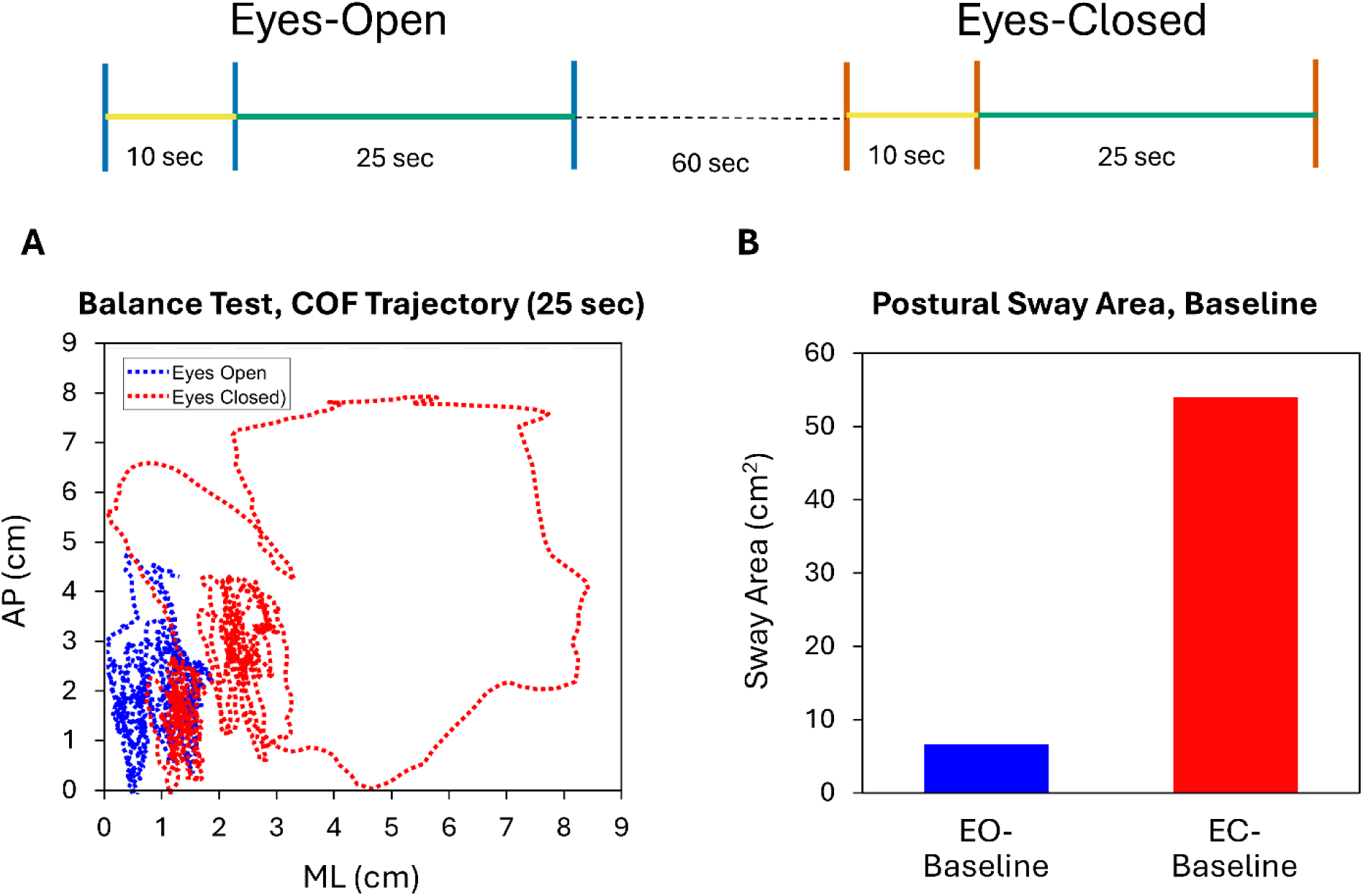
Static balance tests with eyes open (EO) and eyes closed (EC) at baseline visit. (A) Center of force (COF) trajectory sway in the anterior-posterior (AP) and medial-lateral (ML) directions; (B) Postural sway area.

### End of Study Assessments

Both participants tolerated ES lead implantation, task-specific training with ES, and lead explant well with no signs or symptoms of fatigue or MS flares during the clinical trial.

In Participant 1 (specifically the most affected leg), analysis of gait data from the end-of-study assessments with ES-Off revealed notable improvements, including increased hip extension, knee flexion, and ankle dorsiflexion compared to the baseline visit (Figure 7A). The end of study comparison between ES-Off and ES-On conditions (Figures 7B) showed that ES resulted in marked increases in right knee maximum flexion during the swing phase (39%) and hamstring peak muscle activity (30%), a decrease in rectus femoris peak muscle activity (14%), and the restoration of a more natural walking pattern by enabling the foot to go through its typical rolling motion (three rockers of the foot) during each step during the ES-On condition (Figure 8). The gait speed decreased during end of study tests with ES-Off and ES-On in comparison to baseline tests (Table 1). In Participant 2, gait parameters and joint kinematics did not change notably between the ES-On and ES-Off conditions compared to baseline. (Figure 7B-C). Table 1 indicates the spatiotemporal parameters during the baseline assessment and the end of study assessments with ES-Off and ES-On.

**Figure 7.**
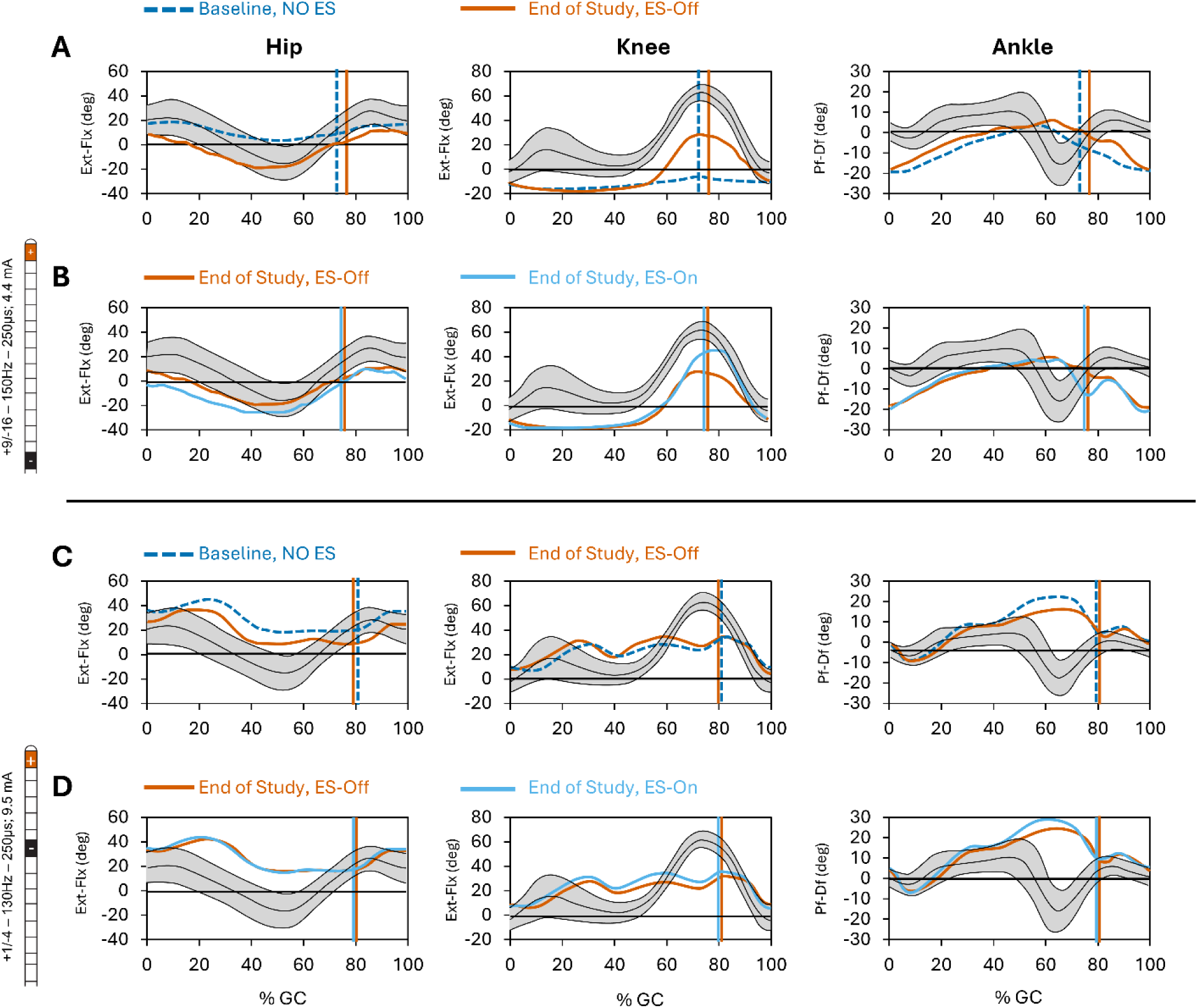
Lower extremity joint kinematics in the sagittal plane are illustrated for Participant 1 (right side) during baseline and ES-Off condition at the end of the study (A), as well as with ES-Off and ES-On conditions at the end of the study (B). Stimulation was delivered using a current amplitude of 4.4 mA, pulse frequency of 150 Hz, and an asymmetric charge-balanced waveform with a pulse width of 250 µs. Similarly, data for Participant 2 (left side) are shown baseline and ES-Off condition at the end of the study (C), as well as with ES-Off and ES-On conditions at the end of the study (D). using a current amplitude of 9.5 mA, pulse frequency of 130 Hz, and an asymmetric charge-balanced waveform with a pulse width of 250 µs. Joint angles for the hip and knee are expressed as extension–flexion (Ext–Flx), and for the ankle as plantarflexion–dorsiflexion (PF–DF). Each plot displays joint angle trajectories over the gait cycle (%GC; 0–100%). Vertical lines indicate the transition from stance to swing phase. Solid black lines represent lab normative data, with shaded regions showing ±1 standard deviation.

**Figure 8.**
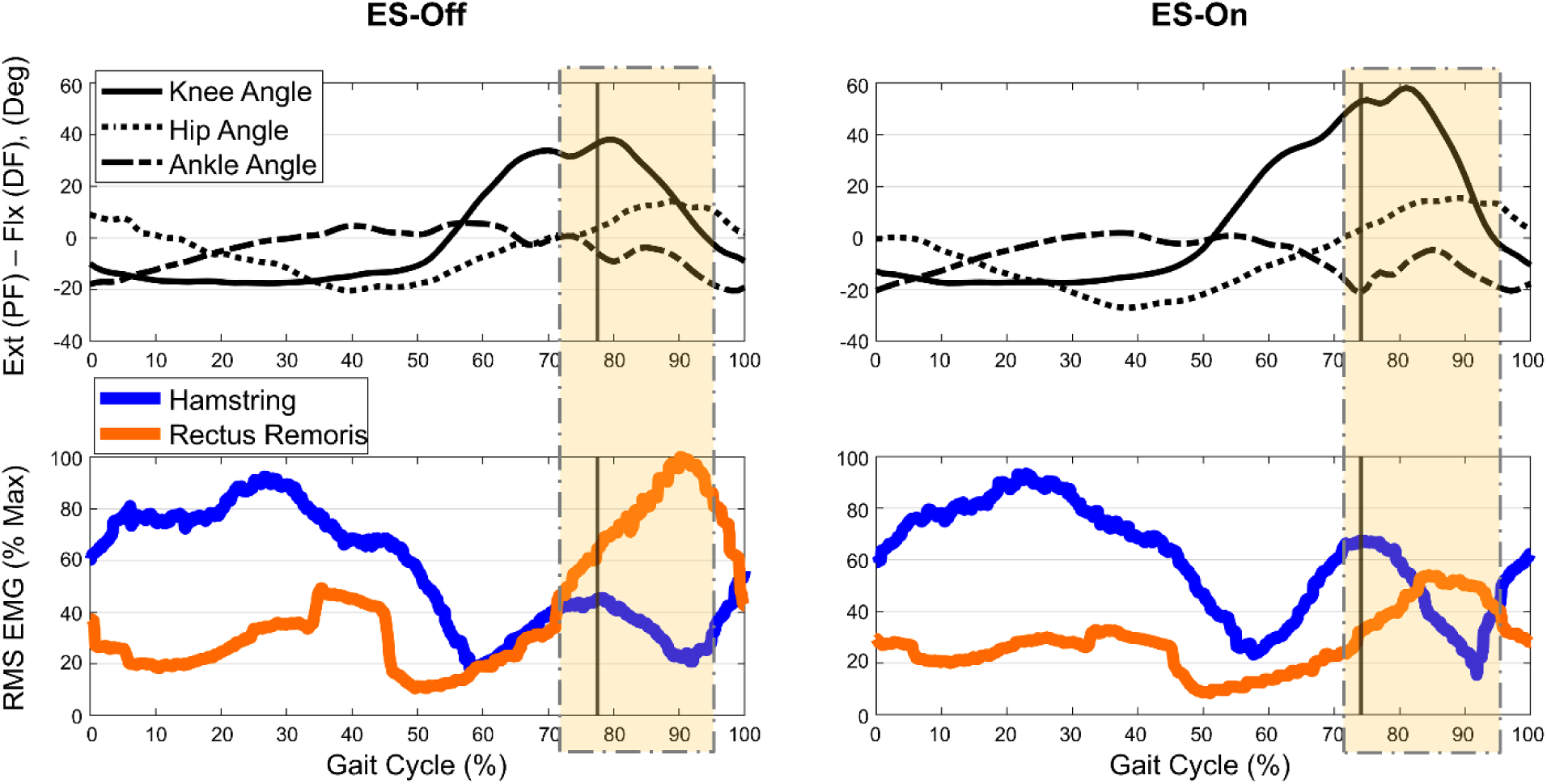
Lower extremity (LE) joint kinematics and Root Mean Square (RMS) (time averaging, period of 0.25 sec) electromyography (EMG) (normalized to the max value during all gait cycles) of hamstring and rectus femoris for a representative gait cycle during post-rehabilitation assessments with ES-Off and ES-On (right limb). Vertical lines represent transition from stance to swing. Highlighted boxes represent the time that kinematic and EMG data were processed (unpublished data).

The MAS score for the right side (Participant 1, most affected leg) knee extensors decreased from 2 during ES-Off to 1^+^ during ES-On (Figure 9), and on the left side (participant 2, most affected leg) decreased from 1^+^ during ES-Off to 1 during ES-On (Table 2). Data from the pendulum test indicated that the first swing angle increased by 6%, and rectus femoris muscle activity decreased by 58% during passive swing of the lower limb with ES-On compared to ES-Off in Participant 1 (Figure 9); however, the pendulum test data were not different between the two conditions for Participant 2 (Table 2).

**Figure 9.**
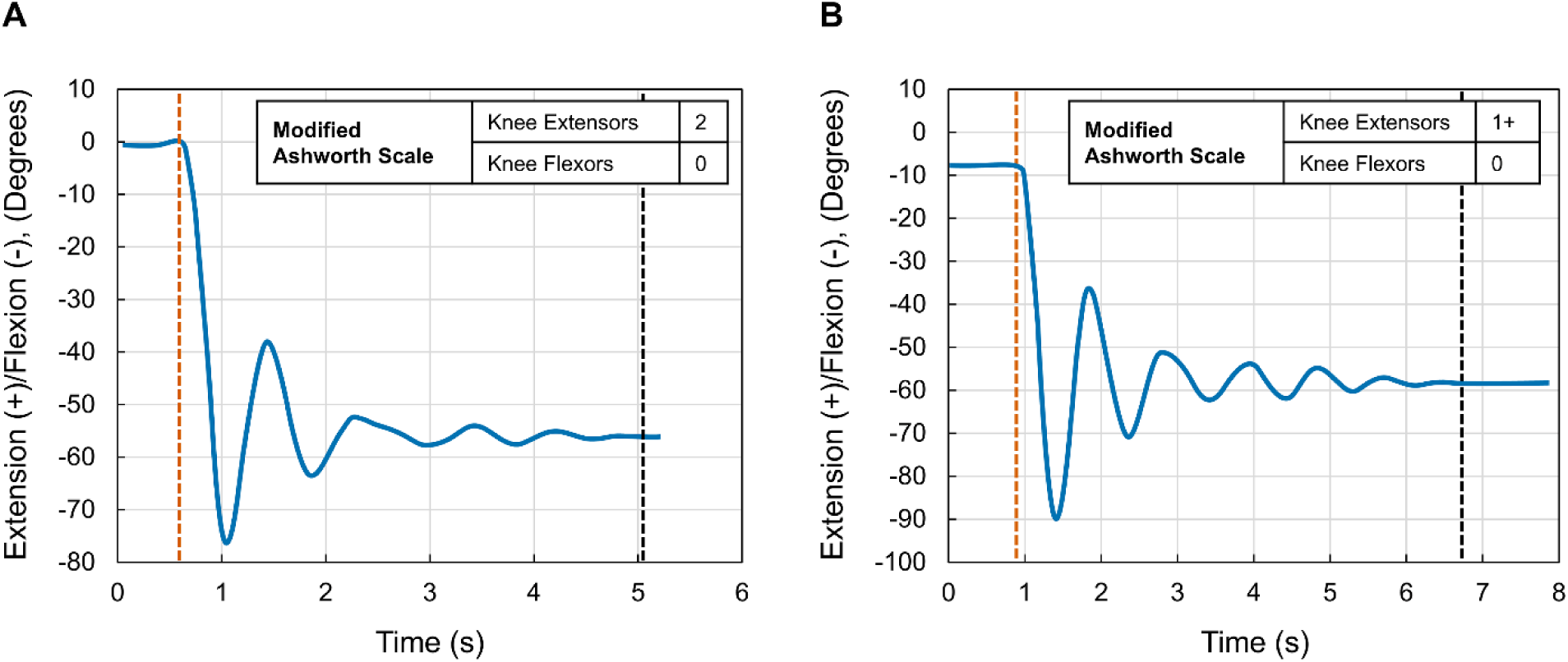
Two representative kinematic recordings during the pendulum test using an IMU sensor : A) in Participant 1 (right side) at the end of study with ES-Off, Modified Ashworth Scale (MAS) = 2, and first swing angle (FSA)= 77 degrees; and B) in the same participant with ES-On, MAS = 1+, and FSA= 91 degrees.

**Table 2.**
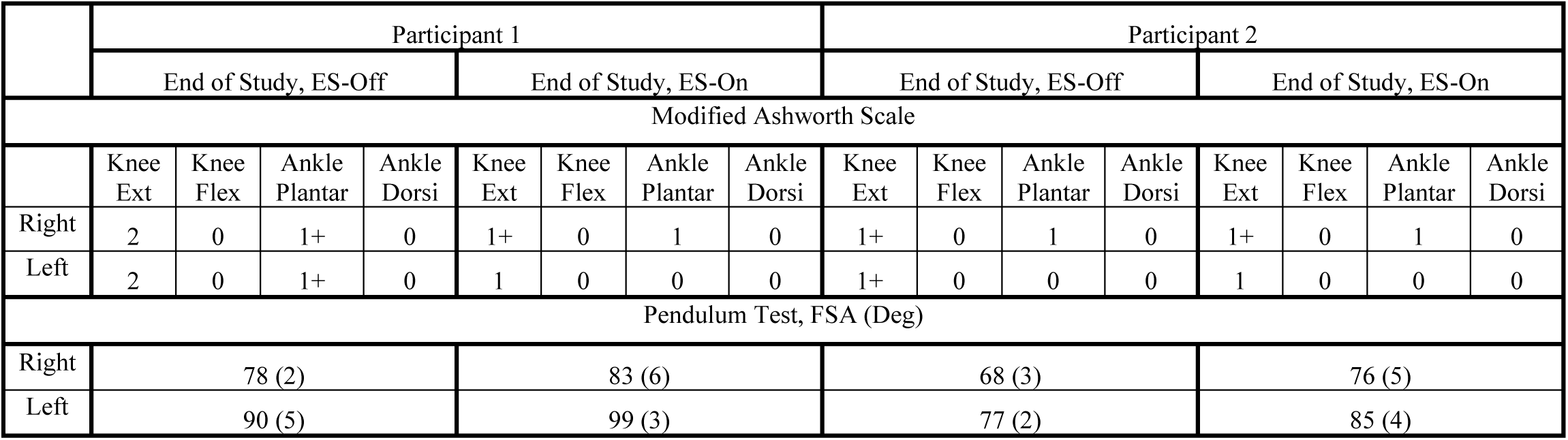
Modified Ashworth Scare (MAS) scores for the knee extensors (Ext), knee flexors (Flex), ankle dorsiflexors (Dorsi), and ankle plantarflexors (Plantar), as well as mean (SD) of first swing angle (FSA) during pendulum test.

|  | Participant 1 |  |  |  |  |  |  |  | Participant 2 |  |  |  |  |  |  |  |
| --- | --- | --- | --- | --- | --- | --- | --- | --- | --- | --- | --- | --- | --- | --- | --- | --- |
|  | End of Study, ES-Off |  |  |  | End of Study, ES-On |  |  |  | End of Study, ES-Off |  |  |  | End of Study, ES-On |  |  |  |
| Modified Ashworth Scale |  |  |  |  |  |  |  |  |  |  |  |  |  |  |  |  |
|  | Knee Ext | Knee Flex | Ankle Plantar | Ankle Dorsi | Knee Ext | Knee Flex | Ankle Plantar | Ankle Dorsi | Knee Ext | Knee Flex | Ankle Plantar | Ankle Dorsi | Knee Ext | Knee Flex | Ankle Plantar | Ankle Dorsi |
| Right | 2 | 0 | 1+ | 0 | 1+ | 0 | 1 | 0 | 1+ | 0 | 1 | 0 | 1+ | 0 | 1 | 0 |
| Left | 2 | 0 | 1+ | 0 | 1 | 0 | 0 | 0 | 1+ | 0 | 0 | 0 | 1 | 0 | 0 | 0 |
| Pendulum Test, FSA (Deg) |  |  |  |  |  |  |  |  |  |  |  |  |  |  |  |  |
| Right | 78 (2) |  |  |  | 83 (6) |  |  |  | 68 (3) |  |  |  | 76 (5) |  |  |  |
| Left | 90 (5) |  |  |  | 99 (3) |  |  |  | 77 (2) |  |  |  | 85 (4) |  |  |  |

In Participant 2, postural sway decreased notably during EC (91%) and EO (50%) conditions at the end of study (ES-Off) compared to baseline. Post-rehabilitation comparison between ES-Off and ES-On conditions demonstrated that ES led to a notable decrease in postural sway during EC (56%) and EO (30%) conditions (Figure 10).

**Figure 10.**
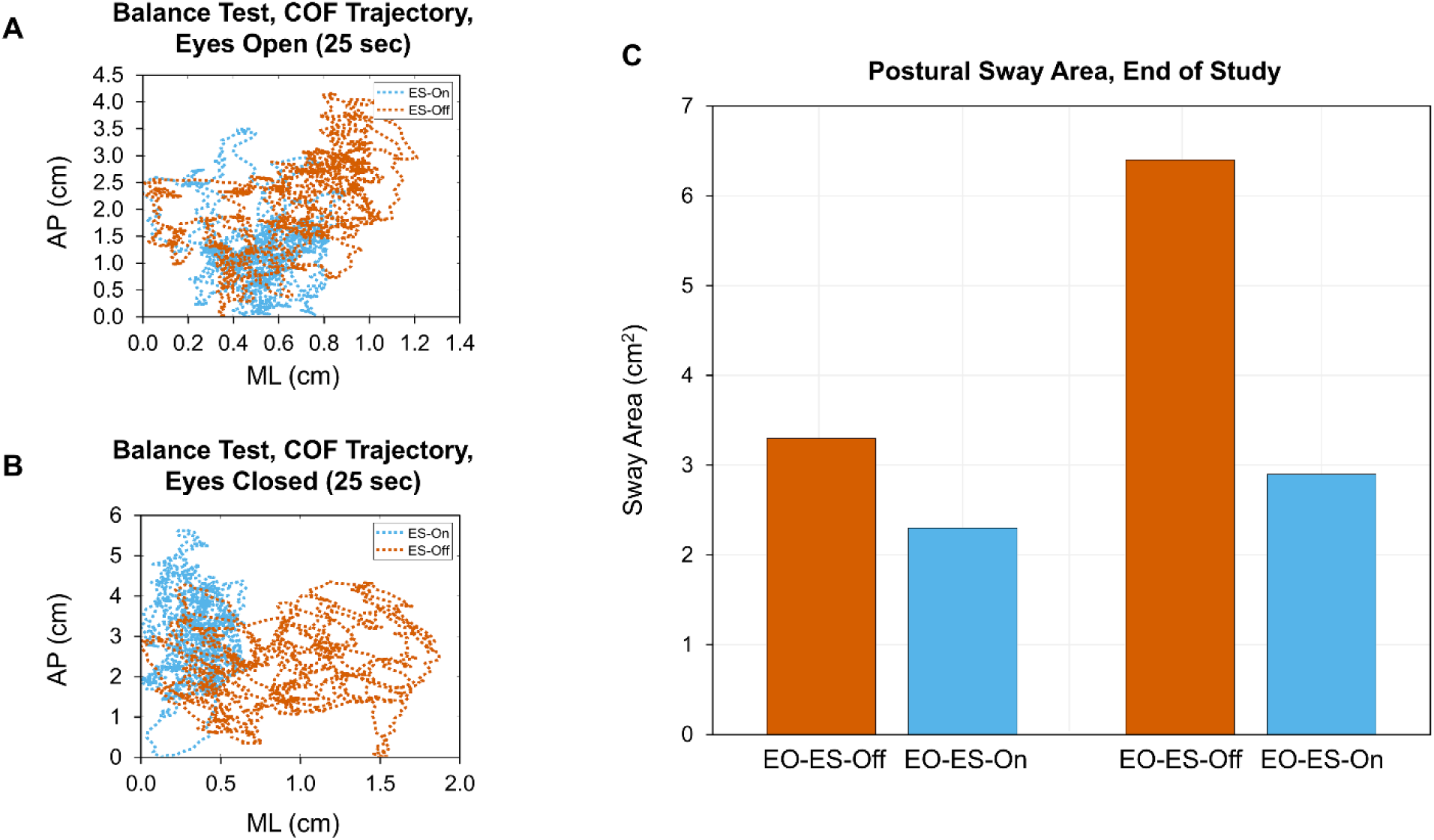
Static balance tests with eyes open (EO) and eyes closed (EC) in Participant 2: The center of force (COF) trajectory sway in the anterior-posterior (AP) and medial-lateral (ML) directions (A & B) and the postural sway area (C) at the end of study.

## DISCUSSION

We present two patients with progressive multiple sclerosis with EDSS of 6.5 who had improvement in gait and balance parameters in response to ES combined with task-specific training. The response to stimulation was different in the two patients, likely due to differences in gait impairments, which are not captured by the EDSS.

### Gait and Spasticity

Participant 1 exhibited a spastic hemiparetic gait pattern characterized by hip and knee flexor weakness and knee extensor spasticity, resulting in a circumduction gait. ES use in this participant led to increased hamstring strength and reduced quadriceps spasticity, ultimately improving gait (Figure 8). These improvements included greater hip extension, knee flexion, and ankle dorsiflexion during the end-of-study assessment (both ES-On and ES-Off) compared to baseline (Figure 7). This suggests a synergistic effect of one month of ES combined with task-specific training on walking ability. The marked increase in right knee maximum flexion during the swing phase, along with increased hamstring peak muscle activity, suggests that ES may enhance volitional motor function during walking. Specifically, it improved hamstring muscle activity while reducing co-contraction of the knee extensors. Additionally, the notable decrease in rectus femoris peak muscle activity may indicate reduced spasticity of the knee extensors during late stance as well as initial and mid-swing (Figure 8). The results from MAS and the pendulum test also support the possible impact of ES on reducing knee extensor muscle spasticity (Table 2).

Participant 2 had greater deficits in strength overall but very little spasticity. ES use in this participant did not result in notable gait improvement. Addressing such leg weakness would require broader and higher-intensity ES, but he was unable to tolerate it.

### Balance

ES use in Participant 2 led to an improvement in static balance with EO and EC (Figure 10). The maintenance of postural control requires the integration of multiple sensorimotor processes (visual, vestibular, and proprioceptive) to generate coordinated movements that maintain the center of mass within the limits of stability. Any adverse alteration in these processes or motor impairments, such as spasticity, results in deficits in postural control and balance. Postural control partly depends on the ability to modulate ankle stiffness,^29, 30^ and individuals with spasticity struggle to adjust their ankle stiffness due to increased reflex modulation, leading to poorer postural control. Sosnoff et al. showed that spasticity significantly contributes to the postural control abnormalities in individuals with MS.^31^ Additionally, ankle joint proprioception is crucial for postural control, as it provides key information about standing postural sway.^32^ In MS, proprioception is impaired due to cerebral or dorsal column involvement,^33^ which may explain the greater postural sway with eyes closed compared to healthy individual.^34^ The results of the static balance tests in this study indicate a notable impact of ES on improving static balance and postural control in Participant 2 who had significant paraparesis. This improvement may be attributed to the reduction in ankle spasticity facilitated by ES; however, the results from MAS did not show any notable changes in ankle spasticity. This might be due to subjectivity of MAS and its poor inter-session reliability.^35^ The pronounced effect of ES observed during the EC condition may be due to its role in enhancing proprioception.

### Limitations and Future Directions

While the small sample size and the absence of a rigorous method for determining effective ES parameters and objectively assessing lower extremity spasticity are the main limitations, this study provides valuable preliminary insights and lays the groundwork for future investigations. Further research should focus on evaluating the impact of ES in a larger sample of individuals with progressive MS to determine effective ES parameters for enhancing gait function.

## CONCLUSION

The preliminary findings from this pilot study suggest that ES, when combined with task-specific training, may have a positive impact on facilitating volitional motor function and enhancing gait in patients with MS. These promising results highlight the need for further research with larger sample sizes to confirm and refine these outcomes, as well as to individualize the application of ES for individuals with MS.

## Author Contributions

OJ contributed to conceptualization (lead), formal analysis (lead), visualization (lead), data collection (support), and writing (lead) of this study; AJA contributed to conceptualization (lead), formal analysis (lead), data collection (support), writing (support), and review and editing (support) of this study; MLG contributed to conceptualization (lead), data collection (lead), writing (support), review and editing (support) of this study; DDV contributed to data collection (support), visualization (support), writing (support), review and editing (support) of this study; KAF contributed to data collection (support), writing (support), review and editing (support) of this study; CAH contributed to data collection (support), writing (support), and review and editing (support) of this study; CJM contributed to project data collection (support), and review and editing (support) of this study; ART contributed to supervision, (lead), review and editing (support) of this study; RS contributed to writing (support), review and editing (support), KRK contributed resources (support), and review and editing (support) to this study; PJG contributed to acquiring resources (support), conceptualization (lead), and review and editing (support) of this study; KDZ contributed resources (lead), funding acquisition (lead), and review and editing (support) to this study; WOT contributed to funding acquisition (lead), conceptualization (support), resources (support), supervision (support), writing (support), and review and editing (lead) to this study.

## Data Availability Statement

The data that support the findings of this study are available from the corresponding author upon reasonable request.

